# Clinical Impact of Ultra-Fast Whole Genome Sequencing in Paediatric Haematology-Oncology Practice

**DOI:** 10.1101/2025.08.29.25334714

**Authors:** Aditi Vedi, Jamie Trotman, Joao ML Dias, Martina Mijuskovic, Sera Choi, Laura Kingham, A Rachel Moore, Sarah M Leiter, Rowena Guermech, Amanda Semerene, Aviva Grisby, Sophie Wool, Victoria Joslin, Arunthethy Mahendrayogam, Jonathan Abrams, Michael Gattens, Esther Blanco, Emmy L Dickens, Charlotte Burns, Matthew J Murray, John A Tadross, Claire Trayers, Catherine E. Hook, Jennifer Becq, Taksina Newington, Ivana Armogida, Zoya Kingsbury, Pascal Grobecker, Ines Vitoriano, Mitch Bekritsky, Jacqui Weir, Louise Fraser, Mark Ross, David Bentley, Matthew Cullen, Sam Behjati, Sean Humphray, David Rowitch, Patrick Tarpey

## Abstract

**Background:** Whole Genome Sequencing (WGS) enhances paediatric cancer diagnosis and management compared with standard molecular assays. However, its clinical utility could be further improved by reducing the National Health Service England (NHSE) turnaround times (TAT).

**Methods:** We evaluated an ‘Ultra-Fast WGS’ (UF-WGS) workflow in a tertiary UK paediatric haematology-oncology unit. Children with suspected or confirmed cancer were recruited over two years (2023-2025), and their tumour, bone marrow and/or blood samples were sequenced on the UF-WGS workflow. All patients underwent concurrent NHSE Genomic Medicine Service (GMS) WGS, serving as the validation benchmark.

**Results:** A total of 54 patients were recruited at diagnosis or relapse. UF-WGS reduced TAT to a mean of 3 days from sample collection, compared with 37 days for GMS-WGS. UF-WGS recalled 95% (143/151) of all clinically actionable somatic and germline variants found by standard NHS GMS-WGS testing. UF-WGS detected an additional 19 clinically actionable variants not found by GMS-WGS. Differences between the two workflows were attributable to tumour heterogeneity in some cases, and low variant allele frequency of those variants identified discrepantly.

Additionally, in 18/35 (51%) prospective cases, UF-WGS enabled demonstrable improvements in care. Clinicians independently judged that 9/19 (47%) of the retrospective cases would have clinically benefited from real-time UF-WGS. UF-WGS provided additional flowcell proximity data, which illustrated the potential to positively impact clinical care.

**Conclusions:** This study indicates the feasibility and utility of UF-WGS and shows added benefits for the clinical management of paediatric cancers, with wider implications beyond this patient group.

**Trial Registration:** 22/WA/0336, NCT 07201038 (clinicaltrials.gov)

## Introduction

The introduction of whole genome sequencing (WGS) has improved the diagnosis and management of paediatric cancers, enabling comprehensive detection of diagnostic, prognostic and therapeutically actionable molecular variants^1–4^. For instance, in childhood acute leukaemia, WGS evaluates cytogenetic risk, agnostic of the clinical and morphological phenotype, to guide treatment decisions^5^. WGS also outperforms standard-of-care (SOC) testing in molecular risk stratification and detection of actionable variants in childhood solid tumours^1,3,6–9^. While the National Health Service England (NHSE) Genomic Medicine Service (GMS) now offers WGS to all children with suspected cancer at diagnosis or relapse, turnaround times (TAT) typically exceed six weeks^10^.

The clinical utility of WGS would in principle benefit from more rapid TAT, particularly where molecular information provides critical opportunities for early intervention, ranging from precision medicine to avoidance of over-medicalisation. There are multiple clinical indications that would benefit from a rapid single molecular diagnostic assay to inform clinical management decisions at the outset. Leukaemia is an exemplar of this, as multiple key diagnostic data can be obtained from WGS, including the cytogenetic profile to determine risk stratification, minimal residual disease (MRD) markers to monitor response and pharmacogenomic profiles to guide timely management.

This study assessed the feasibility and accuracy of a rapid sample-to-results workflow deploying Constellation WGS mapped read technology (Illumina, Inc.; heretofore, Ultra-Fast WGS (UF-WGS)) in a cohort of paediatric haematology/oncology patients at a single NHS tertiary paediatric oncology centre. Our objectives were to determine whether UF-WGS could deliver clinically accurate results within significantly shorter timeframes than GMS-WGS, with the aim of offering meaningful benefits for patient management.

## Results

### Study demographics

Overall, 104 samples from 54 children were analysed via GMS-WGS diagnostic testing and UF-WGS. Thirty-five children with solid and haematological malignancies were offered UF-WGS at presentation (prospective cohort, Figure 1A), selected to broadly represent the range of paediatric haematology and oncology practice. A further 19 patients were selected based on availability of archival tumour DNA and GMS-WGS data with known somatic and/or germline variants of clinical or technical interest (retrospective cohort, Figure 1A). The spectrum of diagnoses encompassed 37 distinct entities, broadly representing the epidemiology of paediatric haematology/oncology in clinical practice (**Figure 1B**). Children with neuroblastoma were under-represented in this study, as small tumour biopsy samples at diagnosis were frequently prioritised for SOC testing in these patients. Children with carcinoma were somewhat over-represented due to the stochastic nature of paediatric oncology presentations, and availability of tumour tissue.

**Figure 1:**
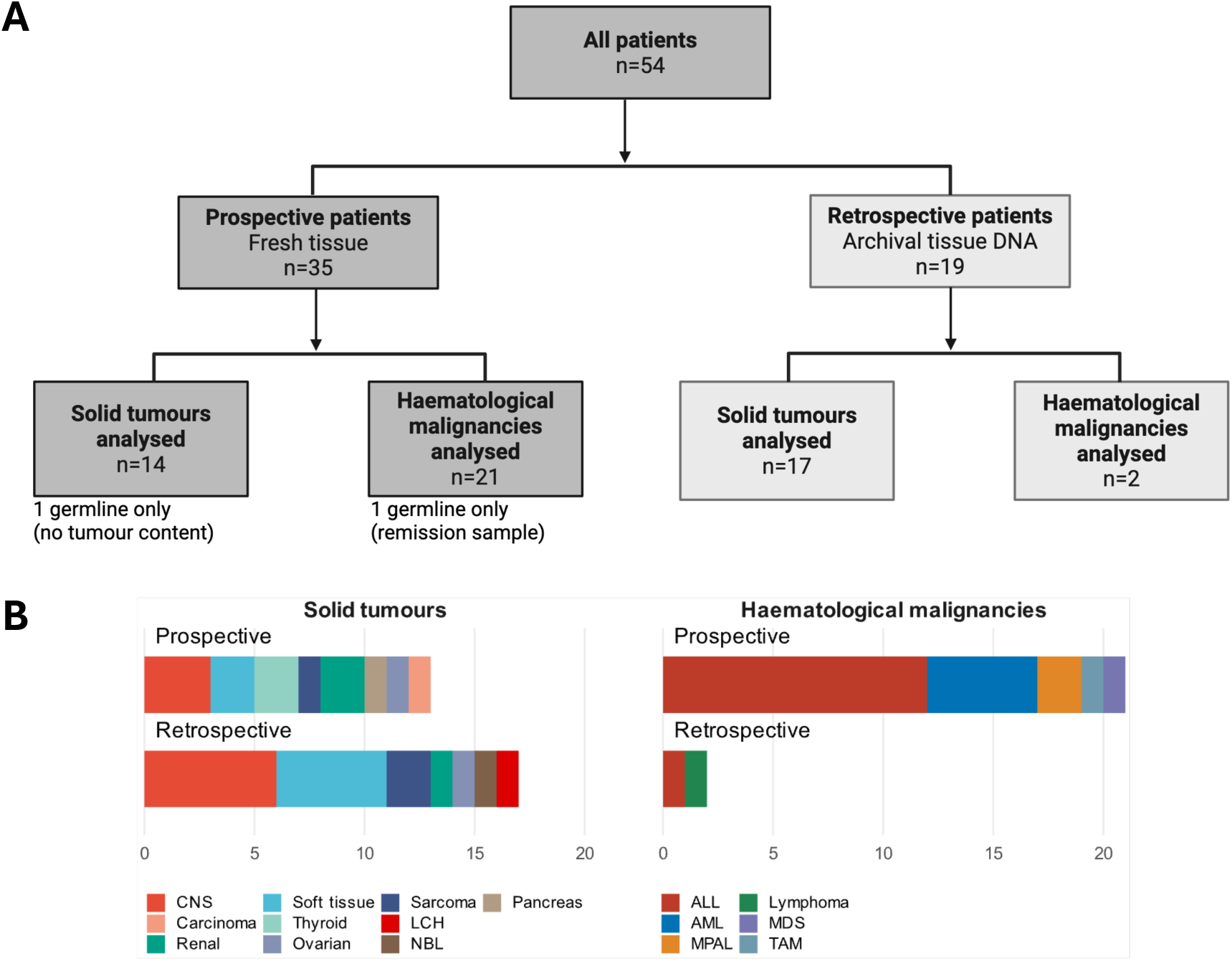
Study design, with 54 patients recruited overall. (A) Prospective patients were recruited at presentation and retrospective patients were selected for availability of archival DNA and GMS-WGS results. (B) Bar plots showing a range of diagnostic groups represented in patients with solid tumours and haematological malignancies in prospective and retrospectively recruited patients. Individual patient information available in Supplementary Table 1. CNS, central nervous system; LCH, Langerhans cell histiocytosis; ALL, acute lymphoblastic leukaemia; AML, acute myeloid leukaemia; MPAL, mixed phenotype acute leukaemia; TAM, transient abnormal myelopoiesis; MDS, myelodysplastic syndrome.

Mean age at diagnosis for the entire cohort was 6.9 years [range 0-17.5 years, standard deviation, (SD) 5.09]. Twenty-three patients (42%) had haematological malignancies, 31 (58%) had solid tumours, with 48 (88%) of patients overall having had samples collected at diagnosis (**Table 1**).

**Table 1:** Study recruitment. (A) Demographics of the overall cohort by solid and haematological malignancies. SD, standard deviation.

| Patient/sample characteristics | Solid Tumours (n=31) | Haematological Malignancies (n=23) |
| --- | --- | --- |
| **Age (years)** |  |  |
| Range | 0-17.5 | 0-15.3 |
| Mean (SD) | 6.6 (5.3) | 7.3 (4.7) |
| **Sex** |  |  |
| Male | 17 (55%) | 17 (74%) |
| Female | 14 (45%) | 6 (26%) |
| **Collection** |  |  |
| Prospective | 14 (45%) | 21 (91%) |
| Retrospective | 17 (55%) | 2 (9%) |
| **Timepoint** |  |  |
| Diagnosis | 29 (94%) | 19 (86%) |
| Relapse | 2 (6%) | 2 (7%) |
| Remission |  | 2 (7%) |
| **Sample** |  |  |
| Paired tumour + germline | 30 (97%) | 2 (9%) |
| Tumour only |  | 20 (87%) |
| Germline only | 1 (3%) | 1 (4%) |

### Comparison with standard molecular testing

UF-WGS captured 95% of somatic and germline variants with clinically actionable diagnostic, prognostic, and therapeutic significance identified by standard molecular diagnostics, including benchmark GMS-WGS. It detected an additional 19 clinically actionable variants not identified by standard assays including GMS-WGS. All variants not detected by UF-WGS were either of low variant allele frequency (VAF), or represented tumour heterogeneity, with different tumour regions being sequenced by each workflow. Non-detection of these variants would not have led to a change in diagnosis or treatment in the clinical context of each patient. UF-WGS returned comprehensive genomic profiling across tumour types, including tumour mutational burden (TMB) and mutational signatures, equivalent to standard GMS-WGS. A visual representation of all clinically actionable variants, TMB and signatures in the cohort is shown in **Figure 2A, 2B, and Supplementary Table 1.** Both sequencing platforms analysed the genome at comparable depths: UF-WGS median depth of coverage was 137x for tumour and 84x for germline samples, GMS-WGS median depth of coverage was 97x for tumour and 42x for germline samples.

**Figure 2:**
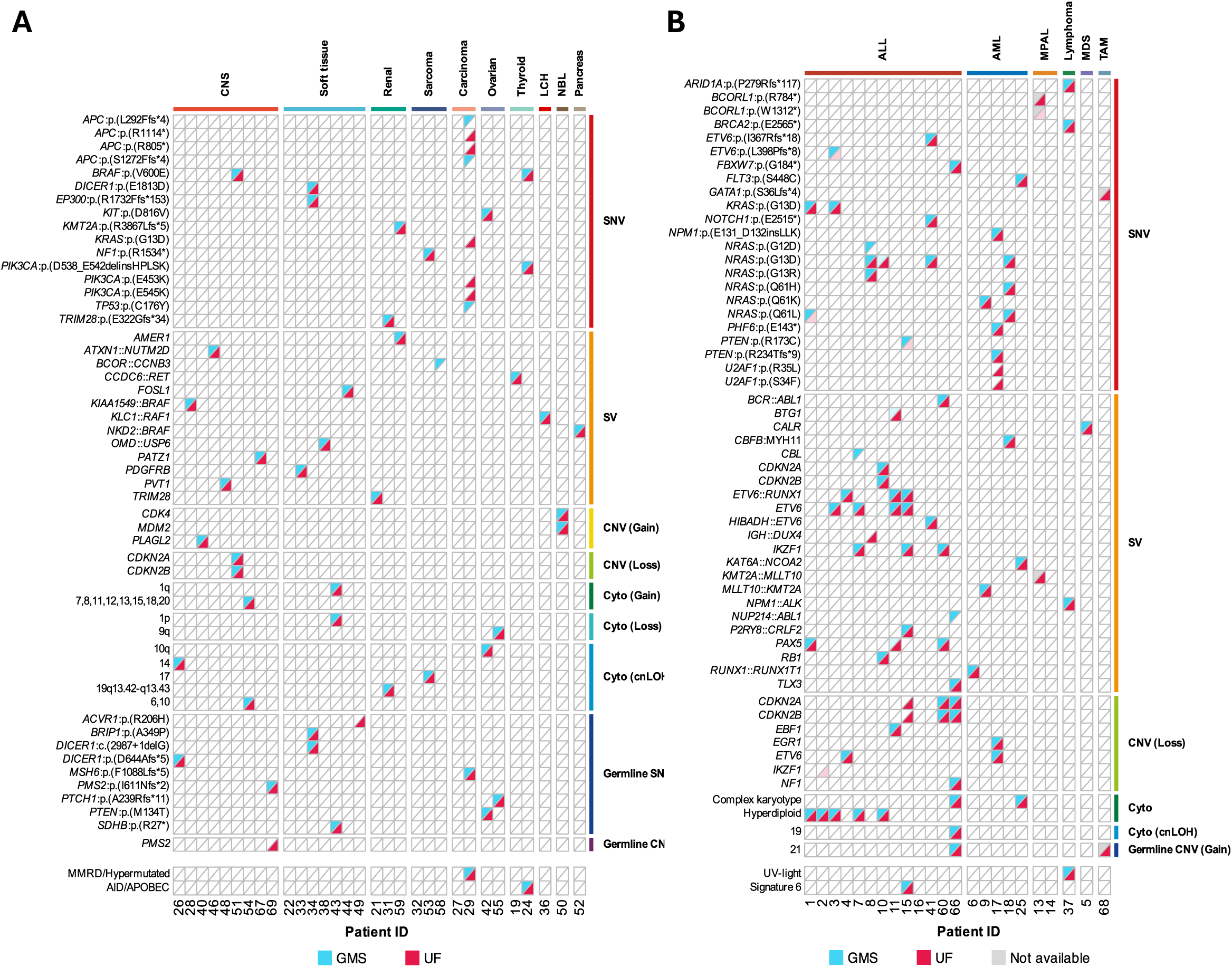
Comparison between GMS and UF-WGS in detection of clinically actionable variants. Oncoplots listing all somatic and germline variants (left) detected, grouped by variant type (right). Only clinically actionable variants as per the National Genomics Test Directory are shown. The cohort is divided into patients with (A) solid (*n=31*) or (B) haematological (*n=*23) malignancies. DNA variants detected by GMS-WGS are shown in red and UF-WGS in blue; cases where DNA quantity was not sufficient for GMS-WGS is indicated in grey. Variants detected at low variant allele frequency are shown for GMS (pale blue) and UF (pink).

There was some expected stochastic variability in calling variants with low VAF <5%, (15 variants in seven patients, **Figure 3A, Supplementary Table 2**). These were more commonly observed in children with leukaemia. In these patients, the final, less tumour enriched bone marrow (BM) aspirate sample was sent for UF-WGS, with more tumour enriched samples being prioritised for standard molecular testing including GMS-WGS. Intratumoural heterogeneity was specifically reflected in the subtle differences in genomic variants identified by the two workflows in a patient with colorectal carcinoma (Patient 29) on a background of congenital mismatch repair deficiency (CMMRD). *KRAS* and *PIK3CA* variants were only detected by UF-WGS, while a *TP53* variant was only detected by GMS-WGS. Different *APC* variants were identified in each of the two assays, without impacting the overall clinical diagnosis. A diagnosis of CMMRD and colorectal carcinoma was independently evidenced by both workflows via detection of a homozygous pathogenic germline *MSH6* variant, elevated TMB and a mutational signature pattern consistent with MMRD. Therefore, the differences in somatic variant detection were not clinically significant. Importantly, the *KRAS* and *PIK3CA* somatic variants detected by UF-WGS are therapeutic targets; both were not identified by GMS-WGS, likely attributable to tumour heterogeneity.

**Figure 3:**
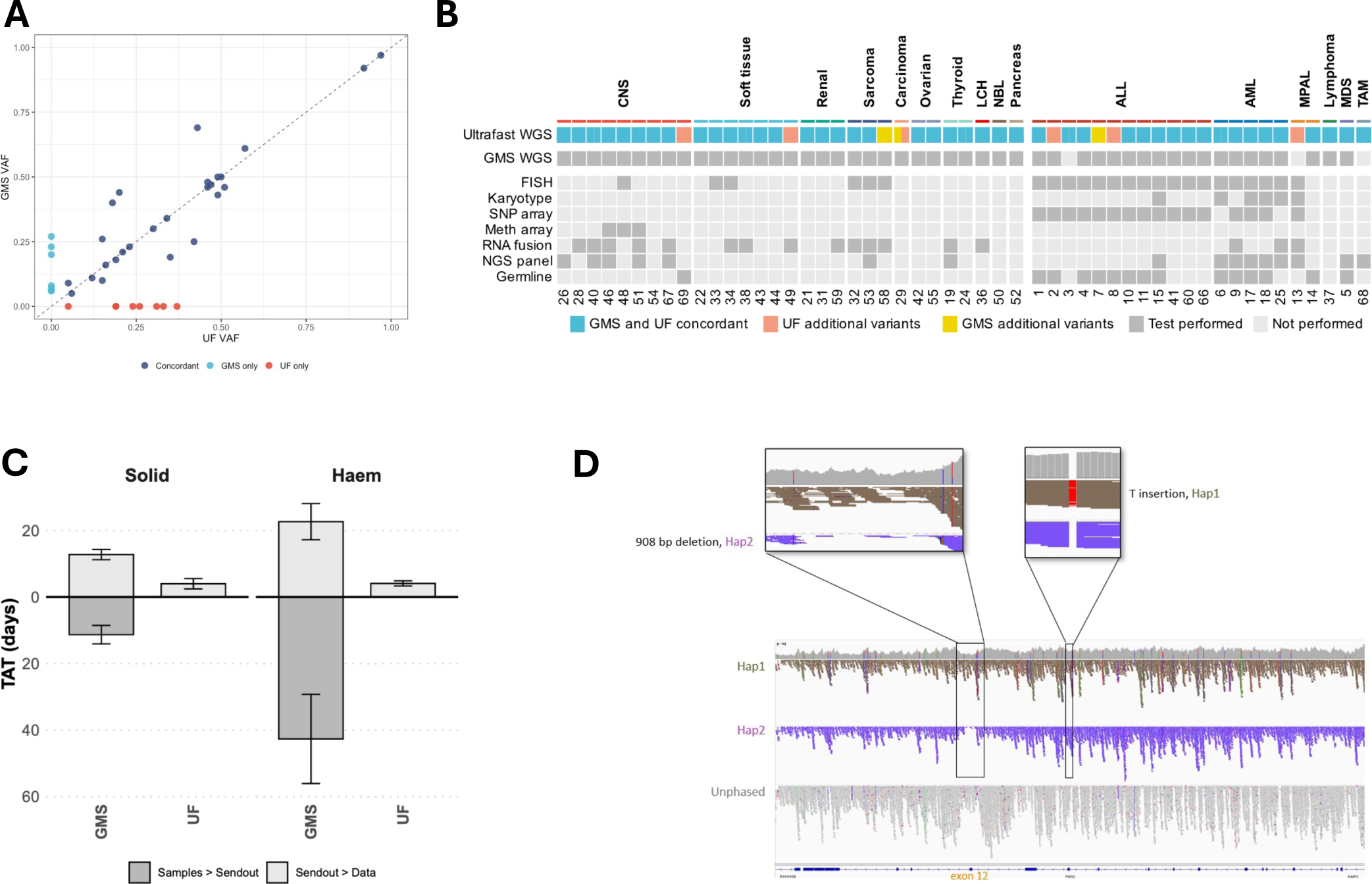
Technical comparison of Ultrafast (UF-WGS) with GMS-WGS and standard-of-care (SOC) assays. (A) Scatter plot representing the variant allele frequency (VAF) for each clinically actionable variant as detected by UF and GMS-WGS pipelines. Only somatic single nucleotide variants (SNVs) or insertions/deletions (Indels) are shown (*n=*41 variants from 19 patients). (B) Heatmap showing SOC molecular testing performed for each patient including GMS-WGS. The top line indicates whether UF-WGS was concordant, discordant or added clinically actionable information compared with all SOC testing, and dark grey boxes indicate which SOC test was performed. Germline testing by SOC was performed using NGS panels when indicated. (C) Bar plots representing median time in days (error bars represent margin of error for a 95% confidence interval) from paired tumour and germline samples (or tumour only for haematological malignancies in UF) being received in the genomics laboratory to send out for sequencing and time to return of complete WGS data in a clinically actionable report format. (D) Identification and phasing of the germline *PMS2* variant in a patient with CMMRD (Patient 69). Phasing of both haplotypes in a difficult to capture region of the genome by UF-WGS. Constellation flowcell proximity data provides long range mapping for phasing of both haplotypes used in the UF-WGS pipeline.

UF-WGS was also concordant with standard molecular testing (not WGS) in detecting clinically actionable variants (**Figure 3B)**, with a faster TAT than all assays, besides fluorescent in situ hybridisation (FISH) in some cases. It provided additional clinically actionable information for six patients above all standard diagnostics, including GMS-WGS (**Supplementary Table 1**).

Clinically interpretable UF-WGS reports were returned within an average of 3 days from sample availability, in contrast to 37 days for GMS-WGS (**Figure 3C**). Differences in TATs are explained in part by delays introduced by skin biopsy and fibroblast culturing required for standard GMS germline analysis in leukaemia. These steps were not required for UF-WGS, which was a tumour-only workflow applying crude lysate from bone marrow (BM)/peripheral blood (PB) or DNA directly onto the flowcell surface.

### Additional insights from UF-WGS technology workflow

The UF-WGS technology provided additional clinically pertinent information. Accurate detection of the *PMS2* gene is challenging via conventional methods including GMS-WGS, related to a number of highly homologous sequences^11–13^. Haplotype phasing and resolving *in cis* versus *in trans* variants within genes of interest was directly beneficial for Patient 69 with a germline *PMS2* frameshift variant and exon 12 deletion, leading to a diagnosis of CMMRD. Phasing confirmed the two *PMS2* variants’ assignment to different parental alleles, differentiating the *PMS2* gene from the *PMS2CL* pseudogene. This would confirm a diagnosis of CMMRD without the need for additional specialised testing (**Figure 3D**). This rapid workflow demonstrates the potential to change clinical practice, allowing clinicians to consider immune checkpoint inhibition for children with CMMRD and avoidance of ionising radiation where feasible from the point of diagnosis.

The additional readouts of proximity mapped data from UF-WGS enhances mapping and clarity of variants which reside in highly repetitive regions of the genome. Using an experimental somatic proximity pipeline in this study, we demonstrated improved sensitivity and specificity of variant calling in such regions, exemplified by the *ATXN1::NUTM2D* fusion in Patient 46, which presented with enhanced clarity as a result of the additional visualisation data from proximity mapping (**Supplementary Figure 1**),

### Clinical impact

We reviewed the potential clinical impact of reduced TAT and additional benefits of the UF-WGS workflow for all patients. Importantly, UF-WGS provided an accurate clinical diagnosis, risk stratification and actionable targets for all 54 patients compared with GMS-WGS and other SOC assays. Additionally, UF-WGS found actionable variants in a patient without cancer (Patient 49), as a result of a disease-agnostic bioinformatic pipeline, which would not have been identified by currently applied GMS assays, leading to a significant change in clinical management. In all cases, UF-WGS findings were orthogonally confirmed by an accredited GMS molecular assay prior to changes in clinical management, in the setting of this being a research-only assay in this study.

In 18/35 (51%) prospectively analysed patients, detection of genomic variants with immediate clinical actionability could lead to an alteration in their clinical management as a direct result of the rapid WGS findings from UF-WGS (**Table 2**). In 9/19 (47%) retrospective cases, three independent clinicians confirmed that UF-WGS results would have altered or improved care pathways had they been available at time of initial presentation (**Supplemetary Table 3**).

**Table 2:**
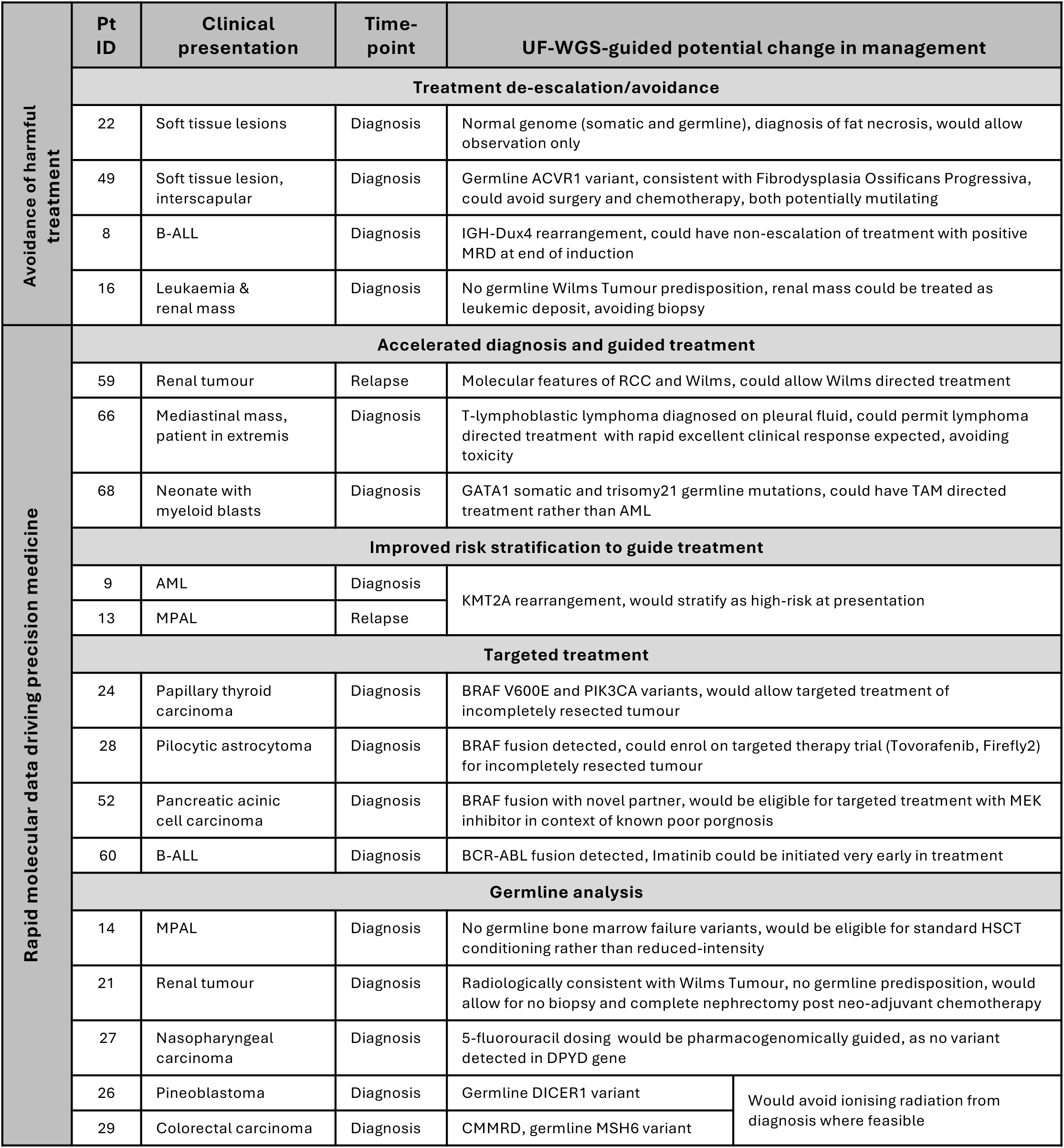
Clinical impact of UF-WGS. List of prospectively analysed patients with a potential change in management guided by rapid molecular information from UF-WGS. B-ALL, B-cell acute lymphoblastic leukaemia; MRD, minimal residual disease; RCC, renal cell carcinoma; TAM, transient abnormal myelopoiesis; AML, acute myeloid leukaemia; MPAL, mixed phenotype acute leukaemia.

#### Case examples

The rapid identification of a novel somatic *NKD2::BRAF* gene fusion, a therapeutically targetable variant, would permit precision treatment for Patient 52 with pancreatic acinic cell carcinoma. This patient would be eligible for targeted treatment with BRAF/MEK pathway inhibitors, within the context of a rare malignancy with known poor prognosis.

Rapid molecular diagnosis would avoid harmful over-medicalisation in several prospective cases. Two patients (22, 49) would be reclassified as having non-malignant conditions, avoiding surgery, chemotherapy and associated toxicities. Patient 22 presented with multiple subcutaneous masses, but had a quiescent genome, which would allow observation to be successfully undertaken with complete spontaneous resolution. Patient 49 had a progressively enlarging interscapular mass, which would be diagnosed rapidly as Fibrodysplasia Ossificans Progressiva (FOP) with pathogenic *ACVR1* germline variant. In this condition, avoiding over-medicalisation is critical because all tissue trauma (e.g., central line insertion) would result in complicating hyper-calcification and irreversible disease progression.

UF-WGS analysis of germline variants helped guide management in three cases. Patient 21 had radiological features consistent with Wilms Tumour, not meeting criteria for biopsy. A germline sample analysed by UF-WGS prior to surgical excision, demonstrated absence of germline cancer predisposition syndromes, which would direct complete nephrectomy with clinical confidence.

Patient 14 with relapsed mixed-phenotype leukaemia (MPAL) could have safely proceed with full myeloablative conditioning after UF-WGS confirmed absence of bone marrow failure (BMF) related germline variants despite subtle clinical features suggestive of underlying BMF, which are diagnostically unreliable^14,15^. Rapid analysis of the *DPYD* gene could have enabled pharmacogenomically guided dosing of 5-fluorouracil in Patient 27 with nasopharyngeal carcinoma^16^.

Patient 66 presented in extremis with a large mediastinal mass and pleural effusion, clinically unfit for a biopsy. A rapid diagnosis of T-cell lymphoma from the pleural effusion would prompt lymphoma directed therapy with rapid clinical improvement, avoiding potential unnecessary toxicity from empiric, non-targeted induction chemotherapy, and invasive biopsy.

### Leukaemia-specific insights

In leukaemia, standard diagnostic pathways require both a BM sample and skin biopsy that is cultured for several weeks, delaying paired tumour/germline WGS analysis. Tumour-only sequencing from BM or PB samples via UF-WGS proved sufficient for identifying clinically actionable mutations, MRD markers, and pharmacogenomic variants. Furthermore, crude lysates used in this study from PB and/or BM samples were directly applied to the UF-WGS workflow, bypassing the need for DNA extraction, enabling even faster TAT.

Patient 13 with relapsed leukaemia was sequenced following a previous haematopoietic stem cell transplant (HSCT). While this compromised the GMS paired WGS diagnostic workflow, UF-WGS was successful with tumour-only analysis to confirm a diagnosis of MPAL, confirmed with SOC molecular assays. Similarly, Patient 68, an infant with a rising myeloid blast count in PB, and a clinical diagnosis of transient abnormal myelopoiesis and was supported by the identification of a somatic *GATA1* mutation within 24 hours via UF-WGS, which would avoid systemic AML therapy, and necessitate appropriate monitoring^17^. UF-WGS also detected the germline trisomy 21 to support the phenotypic features, demonstrating the benefits of rapid tumour-only sequencing even in patients with germline abnormalities.

A subset of 11 patients with paired PB and BM samples showed concordance of variant calls when there were detectable blasts in the PB, supporting the feasibility of using less invasive sampling methods in some patients (**Supplementary Fig 2**).

## Discussion

Our findings demonstrate that UF-WGS is both feasible and accurate in a real-world paediatric haematology-oncology setting, delivering results within 48-72 hours in all cases. Our findings from this pilot study suggest important clinical benefits to time-critical decision making in the diagnosis and treatment of paediatric cancers. Ours is the first study to deploy this technology in cancer, but similar approaches have been applied in paediatric rare genetic diseases^18^.

### UF-WGS outperforms standard-of-care (SOC) testing for solid and haematological childhood cancers

While WGS for paediatric cancer is integrated into national care pathways in England, relatively slower TAT means that it often supplements rather than replaces traditional SOC assays such as FISH, single nucleoptide polymorphisms (SNP) arrays, and targeted panels. Our data suggest that UF-WGS can replace these multiple tests into a single, comprehensive assay with a TAT outperforming SOC testing, enabling clinically meaningful utility of molecular data in diagnosis and management. In leukaemia, for example, the ability to detect MRD markers, cytogenetics for risk stratification including fusion genes (e.g., *BCR::ABL1*, *IGH::DUX4*), and pharmacogenomic variants in a single analysis could both improve and streamline diagnostics. UF-WGS also provides additional insights beyond standard WGS, including genome phasing and improved coverage of difficult genes leading to more accurate variant calling.

### Operational considerations

The highly simplified UF-WGS workflow used in this study is particularly well suited to a decentralised sequencing workflow. High coverage and variant resolution was achieved on a case-by-case-basis without the need for batching samples in contrast to the current centralised NHSE workflow. A rapid and reliable PCR-free genome was delivered for all patients with sufficient tumour and germline DNA content in the samples analysed. Furthermore, tumour-only WGS proved feasible for leukaemia, even in patients with underlying germline variants, consistent with other studies^19,20^. Practically, the ability to apply crude-lysate from BM or PB directly onto the UF-WGS obviated the need for separate DNA extraction or library synthesis, expediting the laboratory workflow.

### Faster WGS TAT benefits patient care

Rapid diagnostics, precision medicine and pharmacogenomics are critical to improving care in paediatric oncology, particularly as we enter an era of treatment de-escalation for low-risk patients. Several cases in this series highlighted the transformative value of UF-WGS with rapid TAT, including non-malignant conditions being able to be spared invasive, potentially mutilating procedures and toxic therapy. Rapid detection of actionable variants can facilitate timely enrolment into clinical trials or implementation of targeted therapies. Also, germline analysis plays a critical role in clinical decision-making. For example, the common scenario of deciding between nephrectomy and nephron-sparing surgery in children with radiologically diagnosed Wilms Tumour can be answered by rapid germline-first WGS, by interrogating the germline for cancer predisposing variants. This approach is widely relevant to all children with renal tumours treated under European guidelines^21^, whereby the surgical approach following neo-adjuvant chemotherapy is currently decided without the knowledge of underlying germline status, since the current GMS workflow mandates paired tumour-germline analysis, and only a small proportion of children are required to have upfront tumour biopsy. Germline findings also enable pharmacogenomic adjustments to reduce treatment toxicity or inform HSCT conditioning regimens.

Although not included in this series, rapid detection of risk-stratifying pertinent somatic variants that alter treatment at the point of diagnosis including *BCR::ABL* in leukaemia, *MYOD1*, *MYCN* amplification, and *BRAF* alteration in solid tumours, are prime examples of the immediate clinical application of a rapid agnostic assay that outperforms most standard molecular diagnostic assays. Risk stratification in neuroblastoma, medulloblastoma, and other childhood cancers increasingly rely on molecular somatic characterisation, with treatment regimens and associated toxicity varying greatly between high and low risk disease. UF-WGS leads to safer, timely treatment, avoiding over-medicalisation. These examples underscore how delayed WGS can result in avoidable morbidity when treatment decisions must proceed in its absence.

### Conclusions

This study demonstrates feasibility and clinical accuracy of UF-WGS in diagnosis and indicates that rapid TAT can improve precision management of paediatric solid and haematological cancers, and avoid harmful over-medicalisation. Indeed, targeted interventions with better outcomes and avoidance of unnecessary treatment, particularly in a patient cohort requiring urgent clinical management, suggests that this technology could lead to safer cancer treatment decisions.

### Limitations

This is a single centre pilot study, which did not deploy a randomised control arm. Therefore, we cannot conclude clinical efficacy of our approach versus SOC, which will require larger multi-centre randomised trials. These are needed to quantify clinical impact, cost-effectiveness and scalable benefits of this approach.

## Supporting information

Raw data

## Data Availability

All data produced in the present work are contained in the manuscript, and supplementary tables. Any additional data may be available upon reasonable request to the authors

https://figshare.com/s/e67fdeaa28a4f9008b48?file=58395373

## Acknowledgements

We thank the patients and families who participated in this study, and Dr Jack Bartram, Dr Iain Kean and Dr Anthony Rogers for comments and useful discussions. Tissue samples were accessed through the Human Tissue Research Bank, which is supported by the NIHR Cambridge Biomedical Research Centre (NIHR203312). This work was supported by the Rosetrees Trust (AV, DHR), Addenbrooke’s Charitable Trust (AV), Isaac Newton Trust (DHR) and the NIHR Cambridge Biomedical Research Centre (NIHR203312). Views expressed are those of the authors and not necessarily those of the NIHR or the Department of Health and Social Care.

## Methods

### Clinical study

This study included prospectively and retrospectively ascertained patients with cancer or suspected cancer aged <25 years in accordance with the NHSE guidelines for clinical-diagnostic WGS. The study protocol was approved by the Health Research Authority and Health Care Research Wales (REC reference 22/WA/0336). Patients were recruited between April 2023 – March 2025, with written informed consent obtained from all participants or their parents/guardians. Sample handling and DNA extraction for all solid tumour and archival samples was carried out in the East Genomics Laboratory Hub, Cambridge University Hospitals NHS Foundation Trust (CUH), Cambridge, UK. All Ultra-Fast whole genome sequencing (UF-WGS) data were generated using the Illumina Constellation technology (Cambridgeshire, UK). All clinically actionable genomic variants identified by UF-WGS were orthogonally confirmed via NHS standard-of-care (SOC) molecular testing prior to changing management.

In the UF-WGS workflow, all prospectively recruited haematological malignancies (except lymphoma) were sequenced directly from peripheral blood (PB) and/or bone marrow (BM) (or pleural fluid for Patient 66) crude lysate in tumour-only mode. Retrospectively recruited haematological malignancies and all solid tumours were sequenced from an extracted DNA sample and analysed using paired tumour and germline DNA samples.

Turn-around time (TAT) was defined in this study as the interval between sample availability for WGS analysis and issuance of a clinically interpretable report. Only clinically actionable variants were reported as defined by the NHSE genomics test directory. All analyses and data visualisation were performed using R Statistical Software (v 2024.12).

### UF-WGS Workflow

#### DNA extraction

Genomic DNA from the peripheral blood and bone marrow was extracted using the QIAmp Blood DNA Mini kit and MagAttract® HMW DNA kit (Qiagen, Germany), following the manufacturer’s protocols. A total of 200µl of peripheral blood and 100µl of bone marrow (diluted with 100µl of PBS) were used as input for the extractions. Genomic DNA from frozen tumour samples was extracted using the MagAttract® HMW DNA kit (Qiagen, Germany), following the manufacturer’s protocols, with a lysis incubation at 56°C for 2 hours. Depending on tissue availability, between 5mg and 20mg of tissue were used as input for the extractions. DNA concentration was determined using the Qubit dsDNA BR Assay kit (ThermoFisher Scientific) and quality was assessed using the genomic DNA ScreenTape assay (Agilent Technologies).

#### Constellation sequencing

Illumina WGS mapped read technology (Constellation) [1] leverages on-flow cell library preparation and uses proximity information from neighbouring nanowells to generate long-range genomic insights using standard SBS sequencing. In this highly simplified method, separate library preparation is eliminated by the use of flow cell-bound transposomes which capture and tagment long molecules of DNA as they are flowed onto the flow cell surface and ensures that adjacent regions in a sample’s genome remain physically proximal on the flow cell.

Clustering and sequencing by synthesis (SBS) are performed as standard, resulting in high quality polymerase chain reaction (PCR) free short-read sequencing data. No modifications to the instrument hardware are required^22^.

The resulting reads from neighbouring clusters can be reconstructed into an interspersed version of the original DNA template molecule, maintaining the accuracy, depth of coverage, and scalability of standard SBS sequencing while adding phasing, enhanced mapping ability, and improved structural variant detection often associated with long-read methods.

#### Alignment and Variant Calling

Sequencing reads were aligned to the linear version of the GRCh38 reference genome. Haematological cancer samples were analysed in the tumour-only mode using DRAGEN somatic pipeline version 4.0, with likely germline small variants called based on their population frequency and gnomAD count > 50 [2]. Solid cancer samples were analysed using paired tumour/normal mode using DRAGEN somatic pipeline version 4.2 [3, 4] and DRAGEN germline pipeline version 4.2 to call germline variants [5]. For all sample types, copy-number variant (CNV) calling was done in the ‘heterogeneous’ mode, allowing subclonal CNV calls. Systematic noise files were used to reduce false positive small and structural variant calls. Only variants with VCF PASS status were included in the downstream analyses and reporting.

#### Tumour Content Estimation

Tumour purity was initially estimated by the DRAGEN pipeline, fitting the purity/ploidy model using B-allele frequencies and coverage data across the whole genome. It was then manually re-estimated using genome-wide plots of B-allele frequencies, coverage levels and somatic small variant allele frequency (VAF) distributions. When the two estimates disagreed, which was sometimes observed in heterogeneous samples, manual estimation was preferred. In cases where DRAGEN pipeline couldn’t provide a confident purity estimate, manual estimate was used as above or by utilizing VAF of the known driver mutation.

#### Annotation and Variant Filtering

Germline and somatic variants were annotated by Nirvana version 3.9.0 [6] using the Ensembl 91 transcript reference database. Pertinent variants, other than pharmacogenomic variants, were triaged via consequence annotations (SO terms): feature elongation, non-synonymous coding sequence variant, unidirectional gene fusion, bidirectional gene fusion, transcript ablation, canonical splice acceptor/donor variants, stop gained, transcript truncation, frameshift variant, stop lost, start lost, transcript amplification, in-frame insertion, in-frame deletion, missense variant, protein altering variant, splice region variant, incomplete terminal codon variant, copy number increase, copy number decrease, copy number change, transcript variant. Somatic and germline structural variants, other than inter-chromosomal translocations, were considered only if they would disrupt the exons of one of the genes or were located within its promoter region.

Pertinent germline and somatic variants were triaged further using gene and genomic region of interest list provided for each cancer and/or variant type **[Supplementary Table 4]**. In the case of no findings, the list was extended to COSMIC cancer gene census v83 (https://www.sanger.ac.uk/data/cancer-gene-census/).

#### Calculation of Tumour Mutational Burden (TMB) and Mutational Signatures

TMB was calculated as the sum of somatic SNVs and indels divided by the size of the GRCh38 genome reference in megabases. Variants with a population frequency >1% in the 1000 Genomes Project were excluded on the basis that they likely represent germline variants.

Mutational signature analysis was performed by calculating the fraction of each somatic mutation type in a specific class and then applying non-negative least squares to decompose the fraction data from each sample into the operative mutational signatures. To calculate fractions, single base substitutions were divided into 96 classes, double base substitutions into 78 classes, and indels into 83 classes using SigProfilerMatrixGenerator [7, 8]. Reference mutational signatures used for decomposition were from the COSMIC 3.3 release [9]. Only signatures supported by >1000 single-base substitutions or >200 indels were considered significant.

#### DUX4r calling

DUX4 rearrangements were called using the customized caller Pelops (feature available in DRAGEN version 4.4+). SRPB threshold of 10 was used to call samples positive for DUX4::IGH, while a threshold of 15 was used for DUX4-other. False positive regions were filtered using a blacklist as described in Grobecker et al [10].

## STAR Methods

### LEAD CONTACT

Further information and requests for resources should be directed to the corresponding author.

### MATERIALS AVAILABILITY

This study did not generate new resources.

### DATA AND CODE AVAILABILITY

The original data files and the R scripts used for analysis and visualization are available from the corresponding author upon reasonable request. All software packages used are publicly available from the CRAN or Bioconductor.

### KEY RESOURCES TABLE

| RESOURCE | SOURCE | IDENTIFIER |
| --- | --- | --- |
| **Deposited Data** |  |  |
| Raw & Processed Data | This study | Available upon request |
| **Software and Algorithms** |  |  |
| R | R-Core Team | v4.5.0 |
| RStudio | Posit | v2025.05.0+496 |
| readxl | CRAN | v1.4.5 |
| dplyr | CRAN | v1.1.4 |
| ggplot2 | CRAN | v3.5.2 |
| ComplexHeatmap | Bioconductor | v2.24.0 |
| circlize | CRAN | v0.4.16 |
| stringr | CRAN | v1.5.1 |
| rvest | CRAN | v1.0.4 |
| reshape2 | CRAN | v1.4.4 |
| grDevices | R-Core Package | v4.5.0 |
| grid | R-Core Package | v4.5.0 |
| Analysis Scripts | This study | See **METHOD DETAILS** |
| **DNA extraction** |  |  |
| QIAmp Blood DNA Mini kit | Qiagen |  |
| MagAttract® HMW DNA kit | Qiagen |  |
| Qubit dsDNA BR Assay kit | Thermofisher |  |
| DNA ScreenTape assay | Agilent Technologies |  |
| **Sequencing data alignment and variant calling** |  |  |
| DRAGEN somatic pipeline version 4.0 |  |  |
| DRAGEN somatic pipeline version 4.2 |  |  |
| DRAGEN germline pipeline 4.2 |  |  |
| Nirvana version 3.9.0 |  |  |

### METHOD DETAILS

#### Analysis 1: Genetic Variant Detection by GMS and UF Platforms

This analysis was performed using two independent Tables in an Excel File (Solid and Haematological cancer types) contains patient data, genetic variants, diagnostic subgroups, and detection across GMS and UF WGS platforms.

- **Environment Setup**: The heatmaps were generated in R script using libraries *readxl*, *stringr*, *rvest*, *ggplot2*, *ComplexHeatmap*, *reshape2*, *dplyr*, *grDevices*, and *circlize*.
- **Data Loading and Preprocessing**: The dataset was loaded from an Excel file. Data was aggregated to create unique entries for each genetic variant per patient. A custom function converted protein change notations in the *Variant* column from three-letter to one-letter amino acid codes. “Yes“/“No” values in the GMS and UF WGS columns were converted to numeric 1/0.
- **Heatmap Visualization**: Two data matrices were created to represent the presence or absence of each variant for each patient on GMS and UF WGS platforms, respectively. A primary heatmap was built using ComplexHeatmap with a custom cell function to render each cell as a rectangle split by a diagonal line. The upper triangle was coloured based on the GMS value, and the lower triangle by the UF value. A second heatmap was created to display the presence of mutational signatures (MMRD/Hypermutated, AID/APOBEC, UV-light, Signature 6). The two heatmaps were combined vertically and the final figure was exported as an SVG file.

#### Analysis 2: Comparative Analysis of WGS and Other Assays

This analysis was performed using an Excel File contains patient data, diagnostic subgroups, and detection across different genomic assays.

- **Environment Setup**: The analysis was conducted in R using the *readxl*, *dplyr*, *ComplexHeatmap*, *circlize*, and *grid* packages.
- **Data Structuring for Visualization**: Data was loaded and converted to a numerical matrix, with patient IDs as row names, the assays detection as columns and missing values imputed to 0. The matrix was transposed so that assays detection were rows and patients were columns. The data was then partitioned into three matrices (Ultrafast WGS data, Routine NHS WGS and all other assays). The top two heatmaps display the detection in Ultrafast WGS and Routine NHS WGS, respectively. The larger heatmap displays all other assays performed in GMS, with rows grouped by assay type. All heatmaps share the same column order, which is determined by the Diagnosis Subgroup and are annotated accordingly. The final figure is a composite of three vertically concatenated heatmaps and was saved as a high-resolution SVG file.

#### Analysis 3: Turnaround Time (TAT) Analysis

This analysis was performed using an Excel file containing turnaround time (TAT) data for Solid and Haematological cancer types and for GMS WGS and UF WGS platforms.

- **Software and Libraries**: The analysis used R with the *readxl*, *dplyr*, and *ggplot2* packages.
- **Data Processing and Visualization**: Data was imported and grouped by Sample Type (Solid and Haem), Platform, and sample processing stage. Summary statistics, including the number of observations, mean TAT, and standard deviation were calculated for each Sample Type. A bar chart was generated using ggplot2 to represent the mean TAT, with error bars indicating the 95% confidence intervals. To visually separate each sample processing stage, the mean values for the “Samples > Sendout” stage were negated on the plot. The final plot was faceted by Sample Type and saved as an SVG file.

#### Analysis 4: Variant Allele Frequency (VAF) Analysis

This analysis was performed using an Excel file containing patient data, diagnostic subgroups, and genetic variant data – including variant type and variant allele frequency (VAF) – for both Solid and Haematological cancer types across the GMS WGS and UF WGS platforms.

- Software and Libraries: The scatter plot was generated using R script with the following libraries: *readxl*, *dplyr,* and *ggplot2*.
- Data Processing and Visualisation: The dataset was imported and filtered to include only somatic single nucleotide variants (SNVs) and insertions/deletions (Indels), based on the “Origin” and “Variant type” columns for both solid and haematological cancer types. VAF values were converted to numeric format and visualized as a scatter plot using ggplot2, with points coloured by concordance between GMS WGS and UF WGS platforms.

### QUANTIFICATION AND STATISTICAL ANALYSIS

#### Heatmap-Based Visual Analysis (Analyses 1 & 2)

These analyses focused primarily on data aggregation and visualization rather than formal statistical testing. The core of the method involved transforming data from a long format into wide-format matrices suitable for heatmaps. This was achieved using functions like dcast or by transposing the data frame. The resulting matrices contained values indicating the detection of a variant in an assay or platform (1 for presence, 0 for absence in Analysis 1, and 1 for UF and GMS concordance, 0.5 for UF additional variants and 0 for GMS additional variants in Analyses 2).

No statistical clustering was performed. Instead, the rows and columns of the heatmaps were explicitly ordered based on predefined factors such as Variant type and Diagnosis Subgroup to facilitate qualitative comparisons between groups and platforms.

#### Statistical Analysis of Turnaround Times (Analysis 3)

For the TAT analysis, 95% confidence intervals (CI) for the mean TAT were calculated for each group using the t-distribution. The formula used was:

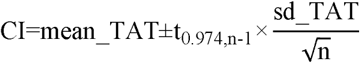

where t_0.975,n−1_ is the critical value from the t-distribution for a 95% confidence level with n−1 degrees of freedom. The calculated means and confidence intervals were then visualized as bar plots with error bars.

