## Supplementary figures for "Clinical Impact of Ultra-Fast Whole Genome Sequencing in Paediatric Haematology-Oncology Practice"

### Supplementary Tables

**Supp Table 1:** All patients in the cohort, including demographics, diagnosis, clinically actionable variants and the platforms by which they were detected.

| Pt ID | Diagnosis | Discrepant variant | VAF | Detected by | Reason for discrepancy |
|---|---|---|---|---|---|
| 29 | Colorectal carcinoma | APC: c.2413C>T; p.(Arg805Ter) | 0.37 | UF | Intratumoural heterogeneity, different slices sequenced by each platform |
| | | APC: c.3340C>T; p.(Arg1114Ter) | 0.33 | | |
| | | PIK3CA: c.1357G>A; p.(Glu453Lys) | 0.24 | | |
| | | PIK3CA: c.1633G>A; p.(Glu545Lys) | 0.31 | | |
| | | KRAS: c.38G>A; p.(Gly13Asp) | 0.26 | | |
| 29 | Colorectal carcinoma | APC: c.875dupT; p.Leu292PhefsTer4 | 0.2 | GMS | |
| | | APC: c.3814dupT; p.Ser1272PhefsTer4 | 0.23 | | |
| | | TP53: c.527G>A; p.Cys176Tyr | 0.27 | | |
| 1 | B-ALL | NRAS: c.182A>T;p.Gln61Leu | 7.00E-02 | GMS | Low VAF, different pulls from bone marrow sample |
| | | ETV6: c.1193_1195delinsCGGAG; p.Leu398ProfsTer8 | 0.06 | GMS | |
| 3 | B-ALL | NRAS: c.35G>A;p.Gly12Asp | 0.06 | GMS | |
| 8 | B-ALL | NRAS: c.38G>A;p.(Gly13Asp) | 0.05 | UF | |
| 10 | B-ALL | PTEN: c.517C>T;p.Arg173Cys | 0.08 | GMS | |
| 15 | B-ALL (relapsed) | U2AF1: c.104G>T;p.Arg35Leu | 0.19 | UF | |
| 17 | AML | U2AF1: c.101C>T;p.Ser34Phe | 0.19 | UF | |

**Supplementary Table 2**: List of the 15 variants from 7 patients that were discordantly identified between UF and GMS. B-ALL, B-cell acute lymphoblastic leukaemia; AML, acute myeloid leukaemia; VAF, variant allele frequency.

**Supp Table 3:** All patients with a potential for change in clinical management as a result of rapid precision molecular diagnostics

**Supp Table 4:** Gene lists applied for prioritisation of variants in the UF-WGS workflow.

### Supplementary Figures

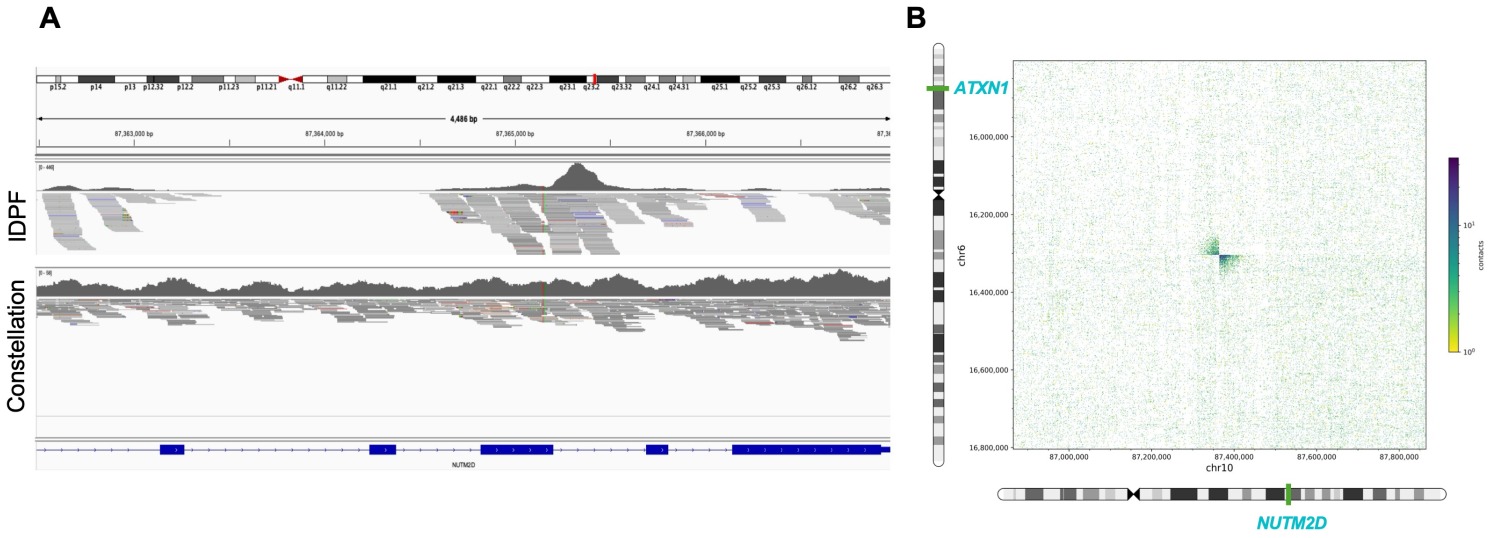


**Supp Fig 1: Additional insights specific to novel Illumina Constellation technology.** (A) Using an experimental somatic proximity pipeline, Constellation closes coverage gaps in high homology *NUTM2D* gene. Reads with mapping quality >20 are shown. IDPF, Illumina DNA PCR-free library preparation protocol (standard). (B) Reciprocal *ATXN1::NUTM2D* fusion in Patient 46 confirmed with long range interactions obtained from flowcell proximity data. Fused regions in both derivative chromosomes are represented as darker-shaded areas of the plot. Breakpoint positions: chr6:16303813-chr10:87364730 (*ATXN1::NUTM2*), chr10:87362944-chr6:16303450 (*NUTM2::ATXN1*).

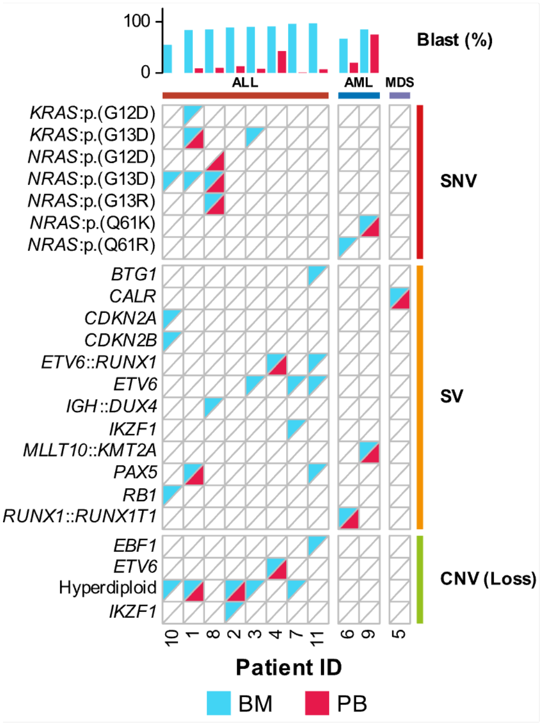


**Supp Fig 2:** Oncoplot showing clinically actionable variants (left) grouped by variant type (right) detected by UF-WGS in the bone marrow (BM) and peripheral blood (PB) from 11 children with haematological malignancies. Blast % is shown in the top bar and is calculated based on flow cytometry of BM samples.
